# Integrative lipidomics and metabolomics for system-level understanding of the metabolic syndrome in long-term treated HIV-infected individuals

**DOI:** 10.1101/2021.05.04.21256640

**Authors:** Sofie Olund Villumsen, Rui Benfeitas, Andreas Dehlbæk Knudsen, Marco Gelpi, Julie Høgh, Magda Teresa Thomsen, Daniel Murray, Henrik Ullum, Ujjwal Neogi, Susanne Dam Nielsen

## Abstract

People living with HIV (PLWH) require life-long anti-retroviral treatment and often present with comorbidities such as metabolic syndrome (MetS). A systematic lipidomic characterization and its association with metabolism is currently missing. In this study, we included 100 PLWH with MetS and 100 without MetS from the Copenhagen comorbidity in HIV infection (COCOMO) cohort to examine whether and how lipidome profiles associated with MetS in PLWH. We combined several standard biostatistical, machine learning, and network analysis techniques to investigate the lipidome systematically and comprehensively. Our observations indicate an increased abundance of the glycerolipids and an association between structural composition patterns of glycerolipids in PLWH with MetS. Further integration of the key metabolites identified earlier in the same population and clinical data with lipidomics suggest disruption of the glutamate and fatty acid metabolism. suggest their involvement in pathogenesis of PLWH with MetS.

## Introduction

Combination antiretroviral therapy (cART) was introduced in 1995 and has increased life expectancy for people living with HIV (PLWH) over the last decades. However, an increase in incidences of comorbidities such as obesity, type 2 diabetes (T2D), and cardiovascular disease (CVD) related to metabolic syndrome (MetS) (i.e., abdominal obesity, hypertension, elevated levels of triglycerides, dyslipidemia, and altered glucose levels), has become a growing concern in successfully treated PLWH. In chronic HIV infection, complicated interactions between effects of persistent low-grade immune activation, metabolic toxicity from cART, and non-HIV related risk factors may increase the risk of MetS in PLWH. However, the pathophysiology of MetS in PLWH is still incompletely understood (Babu et al., 2019; Chai et al., 2019; Gelpi et al., 2020).

cART is known to be associated with changes in fat distribution (lipodystrophy and dyslipidemia) and metabolic abnormalities (Freitas et al., 2011). Changes in fat distribution is a common and known side effect of cART (i.e., affecting up to half of HIV-infected patients receiving cART) and is not limited to a specific drug or active agent. Several studies have investigated the association of HIV infection and the association of cART with metabolic abnormalities related to MetS (e.g., abdominal obesity, T2D, and CVD) (Freitas et al., 2011; Gelpi et al., 2018). These studies have focused on conventional blood lipids, such as triglyceride level and total cholesterol. These biomarkers may not sufficiently reflect the complex alterations of the lipid metabolism in PLWH with MetS. Thus, studies exploring the complexity of the alterations of the lipid metabolism are needed to explore the underlying biomolecular mechanisms of this phenotype.

Untargeted lipidomics is an approach that assesses hundreds of lipid species across multiple biological pathways, which are present in biological samples (i.e., the lipidome). Lipidomics may help in the discovery of new patterns and disease markers associated with MetS in PLWH (Chai et al., 2019). Plasma lipidomics studies in the general population have identified several lipid species within the lipidome to be associated with features of MetS (Meikle and Christopher, 2011). In addition, obesity has been shown to increase the content of almost all detectable diacylglyceride (DAG) and triacylglyceride (TAG) lipid species, along with several cholesterol fatty acids (CE), phosphatidylcholine (PC), phosphatidylethanolamine (PE), and lysophosphatidylcholine (LPC) in a general population (Graessler et al., 2009). The pathophysiology and alterations of the lipidome have yet to be explored. In a prior work from our group, we identified key metabolites, which influenced and altered the metabolome of PLWH with MetS (Gelpi et al., 2021).

An exploratory analysis of the lipidome comparing PLWH without MetS and PLWH with MetS was conducted to identify a set of key lipids that define the mechanism of the lipid abnormalities of MetS in the context of HIV infection. Also, we have performed advanced network analysis that contributed to reveal deeper underlying patterns within the metabolome (i.e., the polar metabolome and lipidome) of PLWH with MetS. Additionally, we investigated the influence of clinical demographic parameters on integrative metabolomics and lipidomics to provide snapshots of the biological phenotypes linked with MetS in PLWH. Our study is the first to provide a comprehensive lipidomics characterization in PLWH with MetS and integrate the lipidomics with a set of key metabolites. The findings can further our understanding of lipid disruption in PLWH with MetS, which may inform future clinical intervention strategies.

## Results

### Machine learning highlights differences in key lipids in PLWH with MetS

PLWH with MetS (n=100) and PLWH without MetS (n=100) were included from the COCOMO study (Table 1). VAT and SAT significantly differed between the two groups (p-value<0.001). The variables immunodeficiency, exposure to early-generation ART, and ART drugs based on their mode of action (i.e., NRTI, NNRTI, PI, INSTI, other/unknown), did not significantly differ (p-value>0.05).

**Table 1:** Clinical and demographic characteristics compared between PLWH without MetS and PLWH with MetS. P-values in bold indicates a significant difference in the concerned variables between the two groups. Immunodeficiency was defined as lowest CD4+ T-cell count <200 cells/*µ*l or previous AIDS condition and exposure to early-generation ART was defined as patients medicated with thymidine analogues, Didanosine and/or Indinavir.

| Variables | PLWH without MetS | PLWH with MetS | pvalue |
| --- | --- | --- | --- |
| Sample (n) | 100 | 100 |  |
| Sex, Male, n (%) | 90 (90.0) | 90 (90.0) | 1.00 ** |
| Age, mean (sd) | 54.4 (9.5) | 54.6 (8.5) | 0.80 * |
| Ethnicity, n (%) |  |  | 0.87 ** |
| Caucasian | 88 (88.0) | 86 (86.0) |  |
| Asian | 3 (3.0) | 2 (2.0) |  |
| Black | 4 (4.0) | 6 (6.0) |  |
| Other/unknown | 5 (5.0) | 6 (6.0) |  |
| Immunodeficiency, n (%) | 14 (14.0) | 13 (13.0) | 1.00 ** |
| Exposure to early-generation ART, n (%) | 34 (34.0) | 46 (46.0) | 0.11 ** |
| VAT, mean (sd) | 76.1 (53.6) | 149.4 (71) | < 1 e-11 * |
| SAT, mean (sd) | 111.1 (71.1) | 150.6 (77.1) | < 0.001 * |
| ART_NRTI, n (%) | 95 (95.0) | 96 (96.0) | 1.00 ** |
| ART_NNRTI, n (%) | 54 (54.0) | 45 (45.0) | 0.26 ** |
| ART_PI, n (%) | 37 (37.0) | 47 (47.0) | 0.20 ** |
| ART_INSTI, n (%) | 16 (16.0) | 21 (21.0) | 0.47 ** |
| ART_other/unknown, n (%) | 0 (0.0) | 3 (3.0) | 0.24 * |

\* Mann-Whitney U test and \*\* Chi-square test

We further applied a number of univariate and machine learning approaches to characterize the effect of MetS in HIV-infected following long-term cART treatment, and to investigate the underlying biological mechanisms of MetS (Figure 1). The lipidomic dataset consisted of 917 unique lipid species including 602 glycerolipids, 228 glycerophospholipids, 61 sphingolipids, and 26 steroids. We observed 618 and 584 significantly differentially abundant lipids between PLWH without MetS and PLWH with MetS (Sup. Data File S1, FDR < 0.001), using Mann-Whitney U and limma, respectively. Moreover, PLS-DA was used to identify variations between the groups based on lipid concentrations, by exploiting its ability to handle a greater number of features compared to samples. Separation of the two groups were indicated by a score plot, where the two first orthogonal components explained half of the variance in the data with 45% and 5%, respectively. We found 516 lipids with VIP values >1, Q2Y = 0.319 (Sup. Data File S1). To obtain a better model performance we could increase the sample size. Finally, we created three types of RF models, minimal-optimal (‘Min’), geometric mean (‘Mid’) and all-relevant (‘Max’) models, which represented feature selection with minimal number of misclassifications, where we observed a good performance of all models (Figure 2a, AUC > 82.9%). Then, we identified 13 lipids as strongest predictors of separating PLWH without MetS and PLWH with MetS (Figure 2b, ‘Max’ MUVR model, AUC > 83%), where the glycerolipid classes, DAGs and TAGs were found to have the greatest significance in group separation.

**Figure 1:**
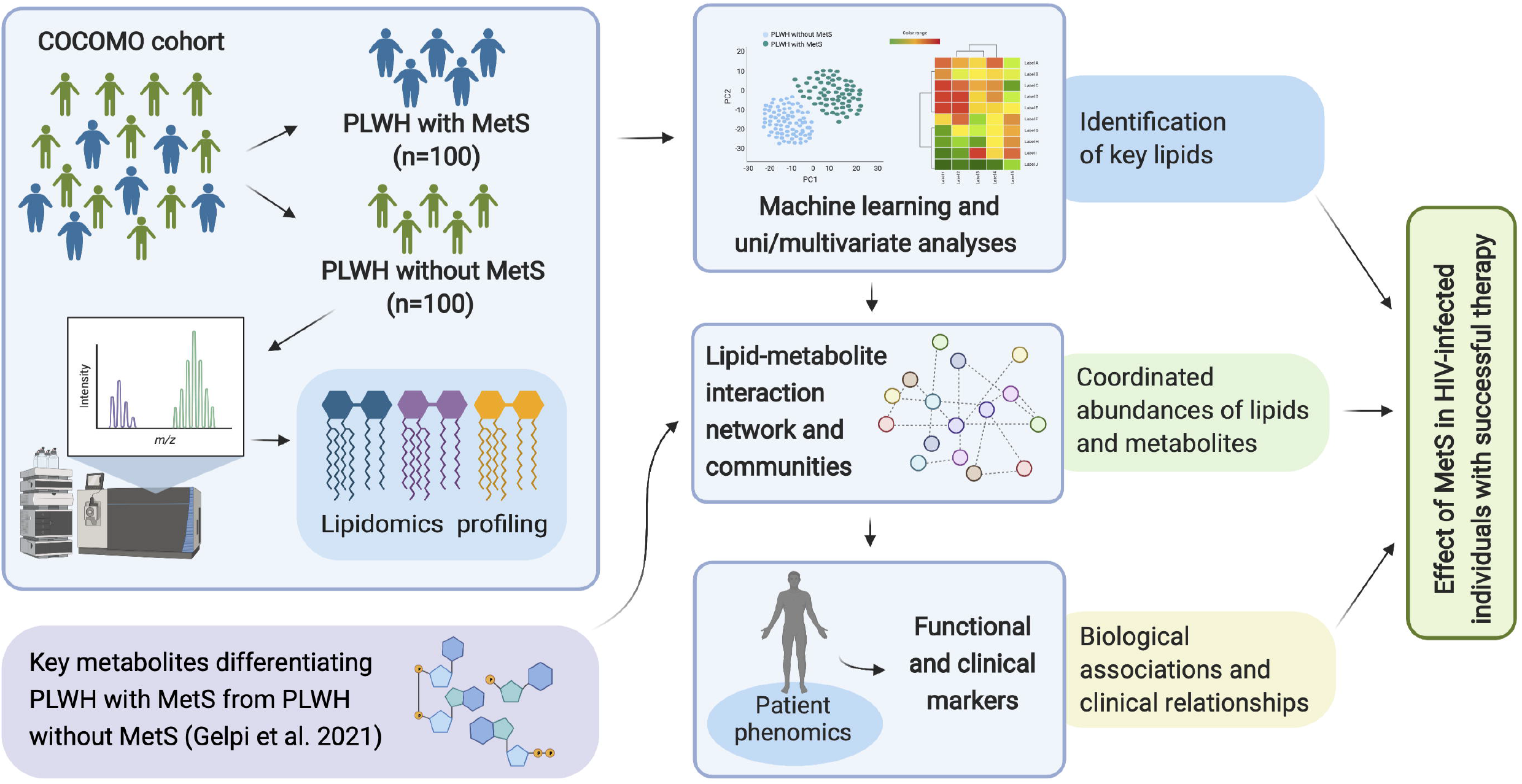
Overview of study workflow. Analysis pipeline for characterizing the effect of MetS in HIV-infected following ART treatment and investigating the underlying biological mechanisms of PLWH with MetS (created with BioRender.com).

**Figure 2:**
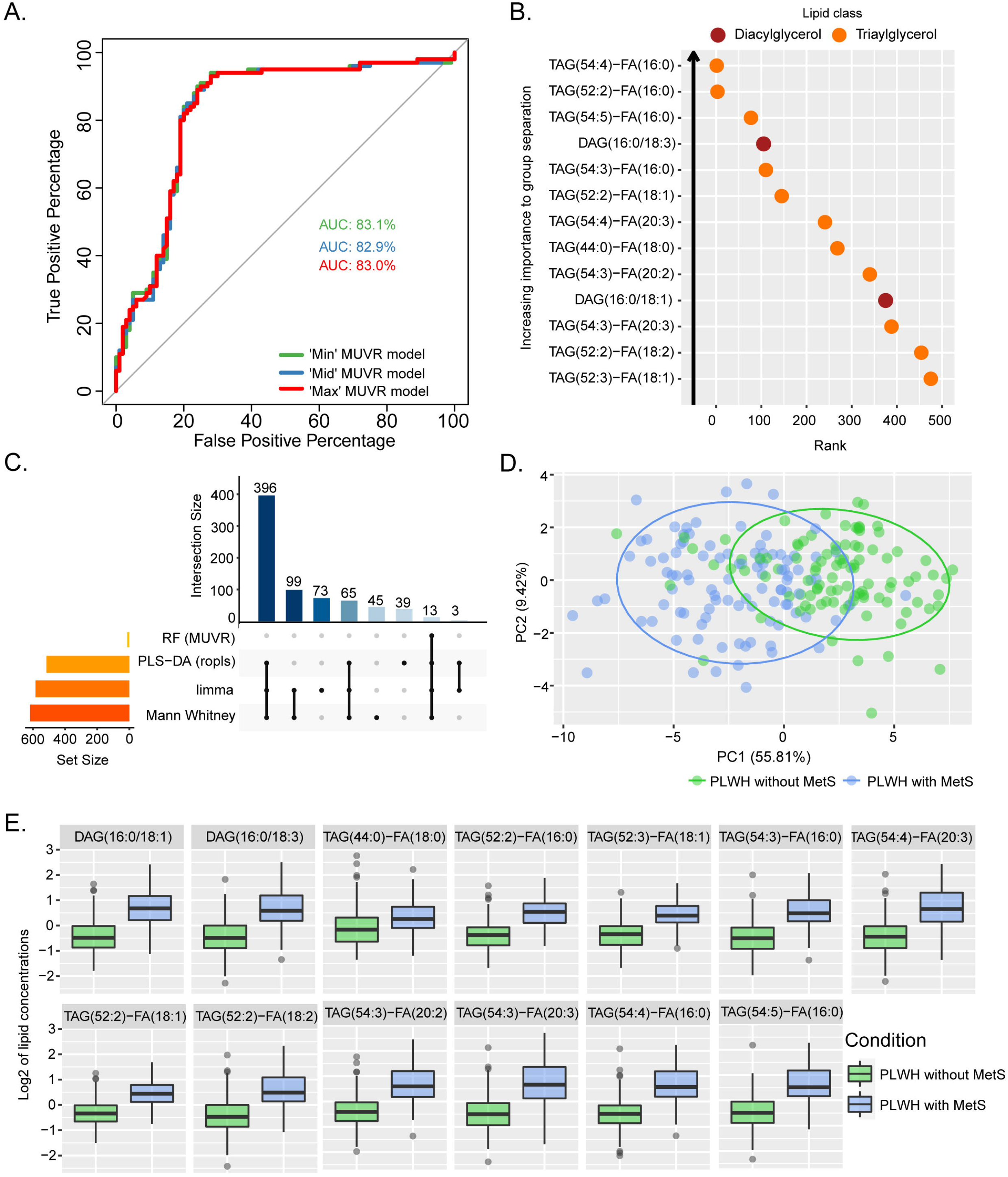
Lipidomics analyses of PLWH without MetS vs PLWH with MetS identifying key lipids differentiating the two groups. (a) Performance of random forest (RF) models. Receiver operating characteristic (ROC) curve with area under the curve (AUC) values for the three *MUVR* models. (b) Important prediction variables separating PLWH without MetS from PLWH with MetS based on lipidomics, diacylglycerol (DAG) and triacylglycerol (TAG). Variables importance on projection (VIP) score plot for the ‘Max’ *MUVR* model, where lower rank indicates better group separation, thus better prediction variables in the model classification. (c) Intersection of methods identifying key lipids. UpSet plot showing number of significant lipids found via four statistical methods (RF, PLS-DA, limma and Mann-Whitney U test). Note the 13 lipids (intersection size on the y-axis) simultaneously identified by all four methods. (d) Separation of PLWH without MetS from PLWH with MetS based on identified key biomolecules. Principal component analysis (PCA) on key biomolecules, where lipidomics and metabolomics data were separated by the 13 identified key lipids and 11 identified key metabolites (Table 2). Ellipses show the 95% confidence interval of the data. (e) Boxplot of lipid concentration of the identified key lipids, which consist of DAGs and TAGs.

The number of significant lipids identified by each of the four methods varied greatly (Sup. Data File S1). However, we observed 13 differentially abundant lipids between PLWH without MetS and PLWH with MetS (Figure 2c) which were consistently identified in all four methods (i.e., Mann-Whitney U, limma, PLS-DA, and RF). These 13 key lipids and 11 key metabolites (Table 2) indicated relatively good separation between PLWH without MetS and PLWH with MetS on sample clustering (Figure 2d). Furthermore, we observed higher abundance level of the key lipids between the groups (Figure 2e).

**Table 2:** Identified key lipids and key metabolites. Overview of key lipids and key metabolites with significant differential abundance between PLWH without MetS and PLWH with MetS. Listed in alphabetical order.

| Key lipids | Key metabolites |
| --- | --- |
| DAG(16:0/18:1) | 1-carboxyethylisoleucine |
| DAG(16:0/18:3) | 4-cholesten-3-one |
| TAG(44:0)-FA(18:0) | 4-hydroxyglutamate |
| TAG(52:2)-FA(16:0) | α-ketoglutarate |
| TAG(52:2)-FA(18:1) | carotene diol (2) |
| TAG(52:2)-FA(18:2) | γ-glutamylglutamate |
| TAG(52:3)-FA(18:1) | glutamate |
| TAG(54:3)-FA(16:0) | glycerate |
| TAG(54:3)-FA(20:2) | isoleucine |
| TAG(54:3)-FA(20:3) | PC/3-MAPC * |
| TAG(54:4)-FA(16:0) | PSP ** |
| TAG(54:4)-FA(20:3) |  |
| TAG(54:5)-FA(16:0) |  |

\* pimeloylcarnitine/3-methyladipoylcarnitine (C7-DC)
\*\* palmitoyl-sphingosine-phosphoethanolamine (d18:1/16:0)

### Structural interpretation of lipids indicates compositional lipid patterns

We then examined the structural characteristics of the lipidome by lipid class in terms of FA carbon number and saturation level (Figure 3). We observed an increase of ceramide (CER), DAG, dihydroceramide (DCER), lysophosphatidylethanolamine (LPE), monoacylglyceride (MAG), PE and TAG, in PLWH with MetS compared to PLWH without MetS, and a decrease in hexosylceramide (HCER) and lactosylceramide (LCER) (Figure 3, FDR < 0.01 and Pearson’s r > 0.7). An increased significantly differential abundance of DAGs and TAGs was observed, indicated by the symbol and red color. The TAGs tended to display a higher abundance of polyunsaturated lipids (i.e., a double-bond content between 2-5) with long-chain fatty acids (LCFA) (i.e., C4856) (Figure 3, FDR < 0.01, Pearson’s r > 0.7). Additionally, TAGs displayed the largest amount of lipid species. DAGs showed a tendency of increase in both saturated and unsaturated lipids (i.e., a double-bond content between 0-6) with LCFA (i.e., C30-40) (Figure 3, FDR<0.01, Pearson’s r>0.7).

**Figure 3:**
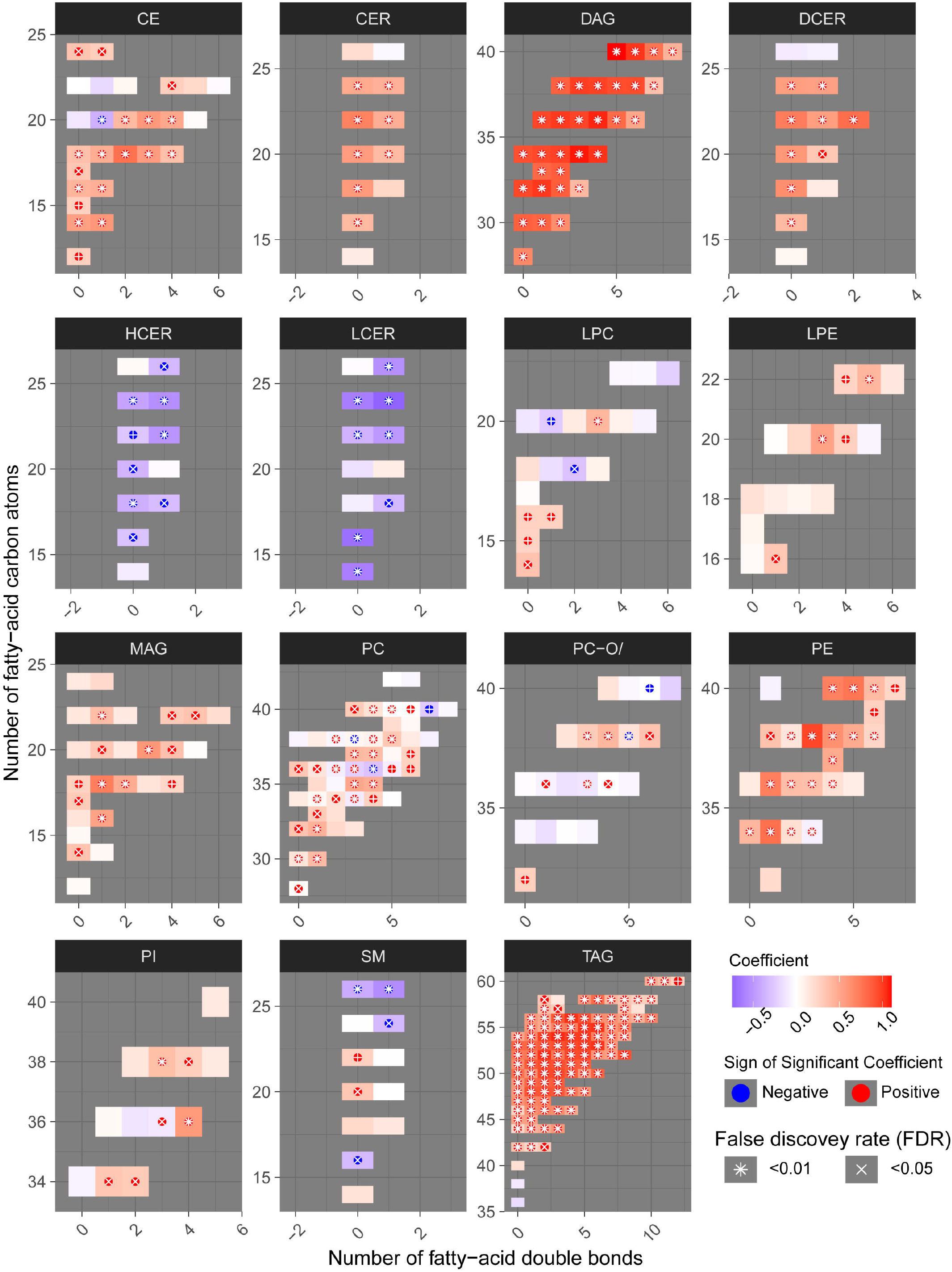
Structural differences of lipidomic profile of PLWH without MetS vs PLWH with MetS. Heatmaps for each lipid class showing the structural lipid composition differences between PLWH without MetS and PLWH with MetS. Each lipid specie is shown as a rectangle and the color shows the abundance difference (red: higher in PLWH with MetS; white: no difference; blue: lower in PLWH with MetS), the lipids were organized by the lipid size (y-axis) and level of saturation (x-axis). Lipids with statistically significant difference between the two groups were highlighted with a symbol. P-values have been FDR adjusted.

### Clinical and omics integrated network identifies biomolecular patterns

Seeking to test whether and how any coordinated patterns of association were present throughout the samples, we generated weighted lipid-metabolite networks. As we aimed to understand the relationship between lipidomic profiles with those metabolites associated with MetS in PLWH. While retaining only informative metabolites we examined the relationship between key metabolites previously identified [11] with the entire lipidome. Briefly, we associated clinical variables with the identified communities and within the most central community we identified associations between clinical variables and each biomolecule.

The fully connected biological network comprised 18430 edges and 917 nodes and displayed markedly distinct behavior from the null network (Sup. Table S1 and Sup. Figure S1). A community analysis on the biological network identified three communities of strongly interconnected lipids and metabolites (Sup. Table S2). Centrality properties were evaluated identifying c1 as the most central community in the network (Figure 4a), which captured most coordinated differential abundance changes. Community c1 had the largest community size (size = 339) and largest community average degree (avg. degree = 534.63) (Sup. Table S2).

**Figure 4:**
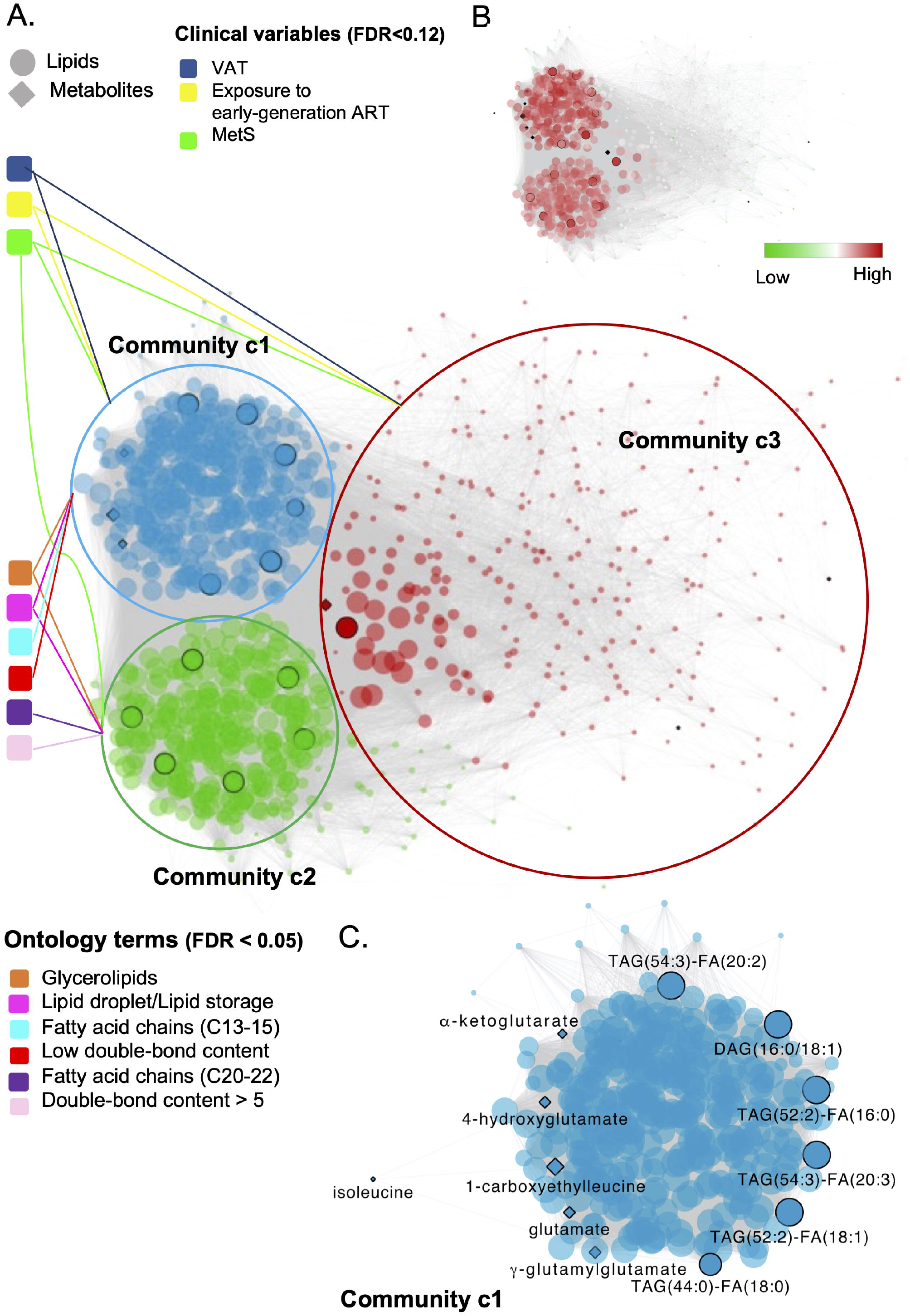
Global and local biomolecular network of PLWH without MetS vs PLWH with MetS. (a) Global network illustrating the associated clinical variables and ontology terms with each community. Network of positive correlations between lipids and metabolites (FDR < 1e-07, Spearman’s ρ > 0.38), colored based on the three identified communities, c1 (blue), c2 (green) and c3 (red). Communities are connected with associated clinical variables (FDR < 0.12) and ontology terms (FDR < 0.05). Black circled lipids and metabolites corresponds to identified key lipids and key metabolites (Table 2). (b) Global network illustrating up and down regulated lipids in PLWH with HIV. (c) Local network of community c1 highlighting key biomolecules. Biomolecular correlations within community c1 (FDR < 1e-07, Spearman’s ρ > 0.38). Black circled and named biomolecules corresponds to the identified key lipids and metabolites within c1.

Structural and functional characterization of these communities [25] (Sup. Figure S2) indicated that glycerolipids and especially TAGs were enriched in both c1 and c2 (Figure 4a, FDR < 0.05). Interestingly, a coordinated structural composition pattern of the glycerolipids displayed average lower carbon number and average lower double-bond content in c1, compared to c2 (Sup. Table S3). Community c3 was not further addressed, as the two other communities were interpreted to be of more importance due to their node-size and average degree (Sup. Table S2). We identified a positive association between community c1 with the clinical variables MetS, VAT and exposure to early-generation ART (Sup. Table S4, FDR < 0.12, illustrated in Figure 4a). In turn, community c2 was positively associated with MetS, however with a lower estimate compared to c1 (Sup. Table S4, FDR < 0.12). Log-fold changes indicated up-regulation of lipids in PLWH with MetS compared to PLWH in both community c1 and c2 (Figure 4b, limma, FDR < 0.001).

Furthermore, we observed a positive association between lipids (DAGs and TAGs) and VAT (Sup. Table S5, FDR < 0.01). TAGs tended to consist of polyunsaturated lipids (i.e., a double-bond content ≥ 2) with LCFA (i.e., C48-54). Interestingly, we also observed that TAGs with LCFA (i.e., C42-48) and a low double-bond content (i.e., ≤ 2) were positively associated with use of NNRTI (Sup. Table S5, FDR < 0.07). Additionally, four out of the 13 key lipids (i.e., TAG(52:2)-FA(16:0), TAG(52:2)-FA(18:1), DAG(16:0/18:1), and TAG(54:3)FA(20:3)) were all found to be independently associated with VAT (Sup. Table S5, FDR < 0.01). Finally, one out of the 11 key metabolites (i.e., glutamate) was also found to be independently associated with VAT (Sup. Table S5, FDR < 0.01).

The top 10% most interconnected biomolecules, found according to their degree, were all glycerolipids within the classes DAG and TAG (Sup. Table S6). Three out of the 13 key lipids (i.e., TAG(52:2)-FA(16:0), TAG(52:2)FA(18:1) and TAG(54:3)-FA(20:3)) were ranked among the top 10% most interconnected biomolecules in c1. Thus, these three lipids were interpreted to be among those biomolecules influencing the behavior of the global network the most. It should be noticed that the structural composition of all three lipids were polyunsaturated TAGs with LCFA and were all found to be positively associated with VAT (Sup. Table S5, FDR < 0.01). No further information was found on the three glycerolipids from the human metabolome data base (HMDB) or KEGG.

The local network of community c1 included six out of the 13 key lipids (i.e., TAG(54:3)-FA(20:2), DAG(16:0/18:1), TAG(52:2)-FA16:0), TAG(54:3)-FA20:3), TAG(52:2-FA(18:1) and TAG(44:0)-FA(18:0)) (Figure 4c). All of the six key lipids within c1 were glycerolipids; five TAGs and one DAG. Interestingly, we observed that all five TAGs were polyunsaturated with a double-bond content between 2 and 4 with a FA carbon number between C52 and C54. Moreover, the key lipids were found to be interconnected with 6 of the 11 key metabolites within c1. Finally, all 13 key lipids and 7 metabolites within the global network were found to be interconnected with each other.

## Discussion

The integrative plasma lipidomics and metabolomics analysis in a large HIV-cohort of PLWH with and without MetS resulted in three main findings that suggests a system-level understanding of MetS in PLWH. First, our data suggested an increased abundance of the glycerolipids DAGs and TAGs in PLWH with MetS. Second, the comprehensive network integration of the lipidomics (analyzed in this study) and metabolomics (previously analyzed (Gelpi et al., 2021)) data suggested interactions between specific glycerolipids structural composition patterns and key metabolites involved in the glutamate metabolism. Finally, our data also indicated a relationship between the structural composition patterns of these specific glycerolipids structural composition patterns with HIV and MetS-specific clinical variables, suggesting their involvement in driving the disease pathogenesis in PLWH with MetS.

In our study, we found 13 key glycerolipids from the classes DAG (n = 2) and TAG (n = 11) to be significantly altered between PLWH with and without MetS. It is worth noting the structural composition of the 13 lipids. The two DAGs [DAG(16:0/18:1) and DAG(16:0/18:3)] consist of unsaturated LCFA (i.e., C34 and 1-3 double-bonds). Additionally, 10 out of the 11 TAGs were polyunsaturated LCFA (i.e., C52-54 and a double-bond content of 2-5). The last TAG had a lower carbon number of C44, compared to the others and was saturated. These findings support previous findings of MetS in general populations that showed that lipids (especially TAGs) with lower carbon number (i.e., C44-54) and lower double-bond content (i.e., 1-4) were associated with an increased risk of T2D. Moreover, it had been observed that an increase in DAGs was associated with hypertension, another MetS-related factor (Hinterwirth et al., 2014). The structure of the FAs is a useful indication of the functionality of the lipid metabolism. Increased accumulation of LCFA such as C(16:0), C(16:1), C(18:0), and C(18:1) suggests increased biosynthesis under MetS-conditions. Such chain compositions are observed among our 13 identified key lipids both in the DAGs and TAGs. Additionally, to the observed pattern of LCFAs, another study suggests LCFAs might cause impairment of mitochondria functions (Hafizi Abu Bakar et al., 2015).

Integrative metabolomics and lipidomics can unravel the complex relationships between metabolites and lipid classes, thus provide a comprehensive view of the metabolic state related to a disease phenotype. We employed network analysis by integrating the key metabolites previously identified as biomarkers in PLWH with MetS (Gelpi et al., 2021) and the lipids with the clinical features (phenomics). Interestingly, we observed that community c1 contained glycerolipids with a lower carbon number and lower double-bond content compared to c2. Community c1 was further investigated and we found that c1 positively associated with the clinical variables MetS, VAT, and exposure to early-generation ART. Our findings are related to previous findings that showed TAGs with a lower carbon number and lower double-bond content play a considerable role in MetS (Rhee et al., 2011; Stegemann et al., 2014). Additionally, our results suggest that exposure to early-generation ART (i.e., thymidine analogues/Didanosine/Indinavir) and increased VAT may also lead to a lower carbon number and double-bond content in glycerolipids, suggesting a role for polyunsaturated glycerolipids with LCFA (i.e., especially TAG(52:2)-FA(16:0), TAG(52:2)-FA(18:1) and TAG(54:3)-FA(20:3)) in the metabolic patterns in PLWH with MetS.

Previous studies have investigated the association of lipidomics profiles (211 lipids) with the progression of CVD in PLWH receiving ART treatment with carotid artery atherosclerosis, compared to HIV-negative individuals (Chai et al., 2019). The study showed elevation in lipid species with polyunsaturated LCFAs (i.e., C13-21 and double-bond content ≥ 2) in patients with atherosclerosis, which is also observed in PLWH with MetS in our study. Additionally, their study suggested significant alterations in lipid species such as cholesteryl ester (CE), LPC, lysophosphatidylethanolamine (LPE), PC, PIs and ceramide (CER). Other studies in both HIV and HIV-negative populations also found alterations in levels of other lipid species (different from DAG and TAG), to be associated with MetS-factors. This includes CE, CER, LPC, PC, PE and sphingomyelin (SM) (Gelpi et al., 2018; Stegemann et al., 2014). Some of these lipid species were also altered in our study population (i.e., CE, CER, PE and SM), however the glycerolipids showed strongest predictive values. Our findings of coordinated abundance shifts in glycerolipids may be due to our considerably larger amount of quantified lipid species (n = 917) compared to other studies (Chai et al., 2019; Rhee et al., 2011; Stegemann et al., 2014) quantified < 215 lipid species each). In the same cluster (c1) we observed two trends with respect to coordinated abundance shifts in glycerolipids. First, TAG species with carbon numbers between C48-54 and double-bond content ≥ 2, together with DAG species with carbon number between C32-36 and double-bond content ≥ 1 associated positively with VAT (FDR < 0.01). This finding correlates with previous studies of MetS factors in HIV-negative cohorts (Rhee et al., 2011; Stegemann et al., 2014). Second, TAG species with carbon numbers between C42-48 and double-bond content ≤ 2 positively associated with the use of ART drugs containing NNRTIs (FDR<0.07). The latter trend supports previous findings suggesting that the NNRTIs drug efavirenz introduce dysfunction in the mitochondria by inducing increased levels of lipids (Blas-García et al., 2010). To our knowledge, we present here the first evidence of association with specific structural composition lipid profiles. Both exposure to early-generation ART and the use of NNRTIs drugs have shown to cause disruption of the mitochondrial functions in previous studies (Blas-García et al., 2010; Trevillyan et al., 2018).

The composition-specific glycerolipids correlated with some of the previously identified key metabolites linked to the perturbations of the glutamate metabolism in PLWH with MetS (Gelpi et al., 2021). This finding correlates with previous studies of MetS in HIV-negative populations, which found branched-chain amino acids (BCAAs) (i.e., leucine, isoleucine and valine) as one of the major metabolite groups dysregulated in obese individuals together with increased concentrations of glutamate, which is the first step of the BCAAs catabolism (Rangel-Huerta et al., 2019; Wang et al., 2020). Additionally, the polar metabolite acylcarnitine (abbreviated PC/3-MAPC, Table 2), an important member of the fatty acid metabolism, was found to be significantly down-regulated in PLWH with MetS. This molecule facilitates the transportation of LCFAs into the mitochondria for catabolism through β-oxidation (Kerner and Hoppel, 2000). Besides glutamate and 4-hydroxyglutamate being a part of the glutamate metabolism, other identified key metabolites were found to be a part of mitochondrial processes, which have important energetic functions (e.g., regulate insulin secretion). These metabolites belonged to the isoleucine metabolism (i.e., 1-carboxyethylleucine, isoleucine) and the TCA cycle (i.e., γ-glutamylglutamate, α-ketoglutarate) (Gelpi et al., 2021).

To our knowledge, correlation between composition specific lipids and polar metabolites has not been seen in previous studies of MetS. In our study the polar metabolites and key lipids are clustered together in the same community (c1) and are positively correlated with each other. These findings suggest a pattern and relationship between polar metabolites involved in the glutamate metabolism and glycerolipids having a specific structural composition of their FA chains (i.e., polyunsaturated LCFAs). To identify this cluster of associated lipids and polar metabolites as potential biomarkers in PLWH with MetS, our findings should be validated in other cohorts.

## Strengths and limitations

Main strengths of this study include a large well-characterized group of PLWH with or without MetS matched on MetS, sex, and age. Furthermore, the use of a fully quantitative lipidomics (i.e., >910 quantified lipid species) methodology allowed us to conduct a thorough analysis of the systematic lipid profiling and its association with metabolites and clinical factors, by using a combination of standard biostatistical, machine learning, and network analysis techniques. The present study also has limitations, such as the cross-sectional design, as no conclusions on causality could be drawn, we were only able to assess the prevalence of the diseases in the plasma samples. Finally, despite the largest study population conducted to date to type comprehensive lipid profile in PLWH, the relatively small sample size of the cohort is also considered as a limitation to this study.

## Conclusion

In conclusion, our study suggests alterations in both the fatty acid metabolism and glutamate metabolism, which both are depending on well-functioning mitochondria. A synergistic effect of different factors (i.e., an increased proinflammatory state induced by HIV, age-related pathophysiological changes, exposure to early-generation ART, and the use of ART with the active agent NNRTIs), which perturb the functions within the biological system of HIV-infected, could play a part in the alterations of the identified biological mechanisms in the phenotype PLWH with MetS. Moreover, our findings also suggest the importance of the structural composition patterns of glycerolipids in the context of excess risk of MetS-related comorbidities among PLWH, such as T2D and CVD. A better knowledge of the structural composition of various glycerolipid species and their association with polar metabolites and clinical variables expands our understanding of the role of lipids in PLWH with MetS. Once we have a better understanding of structural composition patterns of lipid species and their role in biological pathways, novel lipid biomarkers and therapeutic targets could be established to avoid metabolic abnormalities and accelerated aging in PLWH with MetS, as an extension to conventional blood lipid measurements.

## Data Availability

Data and Code Availability:
The lipidomics data datacan be obtained from the dx.doi.org/ 10.6084/m9.figshare.14509452
All the codes are available at github: https://github.com/neogilab/COCOMO_lipidomics

https://figshare.com/articles/dataset/_/14509452

https://figshare.com/articles/dataset/_/14356754

## Acknowledgements

The computations were enabled by resources in project [Dnr. SENS2017550] provided by the Swedish National Infrastructure for Computing (SNIC) at UPPMAX, partially funded by the Swedish Research Council through grant agreement no. 2018-05973. The study is funded by Rigshospitalet Research Council, Danish National Research Foundation (DNRF126) NovoNordisk Foundation. UN acknowledge the support received from Swedish Research Council Grants (2017-01330 and 2018-06156).

## Author contributions

Conceptualization and clinical study designing: S.D.N, S.O.V, U.N, M.G, A.D.K, D.M; Clinical data and biobank: S.D.N., M.G., A.D.K., J.H., M.T.T., and H.U. Methodology: S.O.V., R.B., and U.N., Formal analysis: S.O.V. and R.B., Clinical interpretation: S.O.V, U.N., R.B., S.D.N., M.G., A.D.K. and D.M., Supervision: R.B., A.D.K, M.G., U.N., and S.D.N., Resources: U.N. and S.D.N., Writing (original draft): S.O.V., Writing (review and editing): R.B., A.D.K, M.G., H.U, M.T.T, J.H, D.M, U.N., and S.D.N., Visualization: S.O.V., R.B., and U.N., Project administration: U.N. and S.D.N, Funding acquisition: U.N. and S.D.N. All authors discussed the results, commented, and approved the final version of the manuscript.

## Materials and Methods

### Study designing, patients

We obtained data from the Copenhagen comorbidity in HIV infection (COCOMO) study [12], an ongoing non-interventional, observational, longitudinal cohort study with the aim of assessing the burden of non-AIDS comorbidities in PLWH. Sample collections and quantifications of the COCOMO cohort have previously been described (Gelpi et al., 2018; Ronit et al., 2016). Of the 1099 participants in the COCOMO study, 100 PLWH ≥ 40 years old were included and matched according to age, sex, duration of cART, smoking status and current CD4+ T-cells count to 100 PLWH without MetS (Gelpi et al., 2018; Ronit et al., 2016). MetS was defined as ≥ 3 of the following: (1) waist circumference ≥ 94 cm in men and ≥ 80 cm in women, (2) systolic blood pressure ≥ 130 mm Hg and/or diastolic blood pressure ≥ 85 mm Hg and/or antihypertensive treatment, (3) non-fasting plasma triglyceride level ≥ 1.693 mmol/L, (4) HDL level ≤ 1.036 mmol/L in men or ≤ 0.295 mmol/L in women, and (5) self-reported diabetes and/or antidiabetic treatment and/or plasma glucose level ≥ 11.1 mmol/L (Alberti et al., 2006). For each individual we collected clinical data from the COCOMO database with the following 13 HIV- and MetS specific variables. MetS, sex, age, ethnicity, immunodeficiency (i.e., lowest CD4+ T-cell count <200 cells/*µ*l or previous AIDS condition), exposure to early-generation antiretroviral therapy (ART) (i.e., medicated with thymidine analogues, Didanosine and/or Indinavir), visceral adipose tissue (VAT) [cm2], subcutaneous adipose tissue (SAT) [cm2] and ART drugs including the active agents; nucleotide reverse transcriptase inhibitors (NRTIs), non-nucleotide reverse transcriptase inhibitors (NNRTIs), protease inhibitors (PIs), integrase strand transfer inhibitors (INSTIs), and other/unknown active agents). Furthermore, a lipidomics dataset (see below) and a metabolomics dataset with 11 key metabolites (i.e., 1-carboxyethylisoleucine, 4cholesten-3-one, 4-hydroxyglutamate, α-ketoglutarate, carotene diol (2), γ-glutamylglutamate, glutamate, glycerate, isoleucine, pimeloylcarnitine/3-methyladipoylcarnitine (C7-DC) (PC/3-MAPC), palmitoyl-sphingosinephosphoethanolamine (d18:1/16:0) (PSP)) previously identified by using a combination of standard biostatistical, machine learning and network analysis techniques (Gelpi et al., 2021), were collected. Ethical approval was obtained by the Regional Ethics Committee of Copenhagen (COCOMO: H-15017350). Written informed consent was obtained from all participants.

### Plasma lipidomic profiling

Untargeted lipidomic profiling was performed on plasma samples collected at baseline in COCOMO through the Complex Lipid Panel^*TM*^ technique (Metabolon Inc, Morrisville, NC 27560, USA). Briefly, lipids were extracted from the bio-fluid using automated BUME extraction (Löfgren et al., 2012). Lipids were then transferred to vials for infusion-MS analysis. Samples were analyzed via positive and negative mode electrospray. Lipid species were quantified by taking the ratio of the signal intensity of each target compound to that of its assigned internal standard, then multiplying by the concentration of internal standard added to the sample. Lipid class concentrations were calculated, and fatty acid (FA) compositions were determined by calculating the proportion of each class comprised by summation of individual FAs. All the lipid quantifications were median-centered and missing values were minimum-imputed per lipid species. We further removed variables with zero or near-zero variance from the dataset using nearZeroVar (i.e., 5%, n = 46 of 963).

### Statistics and bioinformatics analysis

All the analyses were carried out in R 4.0.3 (Team, 2016). Clinical characteristics between PLWH without MetS and PLWH with MetS were compared using the Mann–Whitney U test (continuous variables) and chi-square test (categorical variables). Dimension reduction of the key lipids and previously identified key metabolites from the same cohort (Gelpi et al., 2021) were carried out using principal component analysis (PCA). Structural interpretation of the lipidome was carried out through *lipidomeR* (Suvitaival and Legido-Quigley, 2020). We applied different complimentary methods to compare the groups. The normality of the lipidomics data were tested through Kolmogorov-Smirnov test and density plots (Checa et al., 2015). The Mann-Whitney U test was applied to raw data and a subset of lipids with an FDR<0.001, was derived. Log-transformed data were tested for differential abundance using limma and significant lipids with an false discovery rate (FDR)<0.001, were derived (Ritchie et al., 2015). Binary classification modelling was carried out by partial least squares discriminant analysis (PLS-DA) using ropls (Thévenot et al., 2015), where a subset of variables with variables importance on projection (VIP) score >1 was derived. Random forest (RF) was carried out using *MUVR* [https://github.com/CarlBrunius/MUVR], which is developed to create results that are robust with a small sample set. Variables from the optimal RF modelling performance were selected according to rank. Model performance was evaluated by using the Q2Y and area under the receiver operating characteristic (AUROC) for PLS-DA and RF, respectively.

Pathway enrichment was tested from the limma output (FDR < 0.1) with Ingenuity Pathway Analysis (IPA) (Qiagen, US) and MetaboAnalyst (Chong et al., 2019) (limma, FDR< 0.1). The FDR were controlled for by using the Benjamin-Hochberg (BH) method (Checa et al., 2015).

### Network analysis

Network analyses were used to build a biological network consisting of lipids (n = 917) and previously identified key metabolites (n = 11) (Gelpi et al., 2021) after Spearman’s rank correlation across all species. Edges connecting nodes (i.e., biomolecules) were weighted based on positive correlations. This network was compared against a null model attained from a random network with the same number of nodes and edges based on the Erdos-Renyi model (Barabási and Oltvai, 2004). All networks were built through the Python module igraph.(Csardi and Nepusz, 2006) Communities within the biological network were detected through the Leiden algorithm (Traag et al., 2019). Communities were characterized functionally and phenotypically through the lipid specific ontology web-tool, LION/web (Molenaar et al., 2019). LION/web was used to determine lipid ontology trends within each community, using all lipids from the network as background list. Separate analyses on each network community with all lipids as background list was uploaded to LIPEA to identify lipid pathway enrichment (Acevedo et al., 2018). Community association with clinical parameters was determined through logistic and linear regression in R. Network visualization was performed using Cytoscape 3.5.1 (Shannon et al., 2003).

## Data and Code Availability

The lipidomics data datacan be obtained from the dx.doi.org/10.6084/m9.figshare.14509452 and metabolomics dx.doi.org/10.6084/m9.figshare.14356754

All the codes are available at github: https://github.com/neogilab/COCOMO_lipidomics

## Supplementary Files

**Supplementary Data File S1:** Overview of statistical outcomes from Mann Whitney U test, limma, PLS-DA and random forest. Outcome from statistical and machine learning methods identifying lipid species differentiating PLWH without MetS from PLWH with MetS. The table includes significant lipid species and their super pathway together with results from the four used methods Mann Whitney u test (pvalue and FDR adjusted pvalues), limma (pvalue and FDR adjusted pvalues), PLS-DA (variable importance on projection (VIP) values) and random forest (lowest rank indicating strongest predictors of separating PLWH without MetS from PLWH with MetS). Cells with hyphens illustrates that the concerned lipid specie is found not to significantly differentiating the two groups by the concerned method.

**Supplementary Table S1:** Network properties of the positive and random network. Including type of network (Network), node count (Nodes), edge count (Edges), average degree of network (AvgD), average path length (AvgPL), clustering coefficient (CC), if the network is connected or not (C?) and the minimum cut off (MinCut).

**Supplementary Table S2:** Table of community properties including size and average degree of the three identified communities. The size corresponds to the number of lipids and metabolites (nodes) within each community. The average degree of the community corresponds to how connected lipids and metabolites (nodes) within the concerned community are.

**Supplementary Table S3:** A structural composition table, providing an overview of composition of enriched glycerolipids in the network communities c1 and c2.

**Supplementary Table S4:** Correlation table predicting clinicalvariables based on the community score (FDR < 0.12). Ranked according to FDR (p-value adj).

**Supplementary Table S5:** Association table predicting clinical variables based on each lipid and metabolite concentration (FDR < 0.07) within community c1. The model is adjusted for MetS, sex and age. Ranked according to FDR (p-value adj). Five key lipids and one key metabolite were associated with VAT and marked in bold in the table.

**Supplementary Table S6:** Top 10% nodes in community c1 based on the degree. Thus, the most interconnected nodes in the community. The lipids TAG(52:2)-FA(16:0), TAG(52:2)-FA(18:1) and TAG(54:3)-FA(20:3) are marked in bold, as they were among the key lipids.

**Supplementary Figure S1:** Degree distribution for the positive weighted against the random network.

**Supplementary Figure S2:** Ontology enrichment plots for the three communities c1, c2 and c3. (a) Ontology enrichment plots for community c1 (blue) and c2 (green). (b) Ontology enrichment plot for community c3.

