## Supplementary Table S1-S6 and Fig S1-S2 for "Integrative lipidomics and metabolomics for system-level understanding of the metabolic syndrome in long-term treated HIV-infected individuals"

Supplementary Tables

**Supplementary Table S1:** Network properties of the positive and random network. Including type of network (Network), node count (Nodes), edge count (Edges), average degree of network (AvgD), average path length (AvgPL), clustering coefficient (CC), if the network is connected or not (C?) and the minimum cut off (MinCut).

| Network | Nodes | Edges | AvgD | AvgPL | CC | C? | MinCut |
| --- | --- | --- | --- | --- | --- | --- | --- |
| Positive weighted | 917 | 184304 | 402 | 1.72 | 0.87 | True | 1 |
| Random | 917 | 184304 | 402 | 1.56 | 0.44 | True | 351 |

**Supplementary Table S2:** Table of community properties including size and average degree of the three identified communities. The size corresponds to the number of lipids and metabolites (nodes) within each community. The average degree of the community corresponds to how connected lipids and metabolites (nodes) within the concerned community are.

| Community | Biomolecules | Avg. degree | Lipids | Metabolites |
| --- | --- | --- | --- | --- |
| c1 | 339 | 534.63 | 333 | 6 |
| c2 | 318 | 506.58 | 318 | 0 |
| c3 | 260 | 101.05 | 257 | 3 |

| Communities | c1 | c2 |
| --- | --- | --- |
| Biomolecules in community | 339 | 264 |
| TAGs in community (n, %) | 252 (74%) | 318 (83%) |
| Average TAG carbon number | 49 (38-56) | 55 (48-60) |
| Average TAG double-bond content | 2 (0-8) | 5 (0-12) |
| DAGs in community (n, %) | 28 (8%) | 24 (9%) |
| Average DAG carbon number | 32.5 (28-36) | 37 (34-40) |
| Average DAG double-bond content | 1.5 (0-5) | 5 (2-8) |

### **Supplementary Table S3:** A structural composition table, providing an overview of composition of enriched glycerolipids in the network communities c1 and c2.

**Supplementary Table S4:** Correlation table predicting clinical variables based on the community score (FDR < 0.12). Ranked according to FDR (p-value adj).

| Community | Clinical variable | estimate | std. error | t value | p-value | p-value adj |
| --- | --- | --- | --- | --- | --- | --- |
| c1 | MetS | 0.29 | 0.04 | 7.31 | 6.48e-12 | < 0.000001 |
| c3 | MetS | 0.31 | 0.04 | 7.30 | 6.73e-12 | < 0.000001 |
| c1 | VAT | 36.34 | 6.07 | 5.99 | 9.83e-09 | < 0.000001 |
| c3 | VAT | 42.96 | 6.15 | 6.98 | 4.31e-11 | < 0.000001 |
| c3 | Exposure preART | 0.13 | 0.05 | 2.86 | 0.004 | < 0.12 |
| c2 | MetS | 0.19 | 0.08 | 2.32 | 0.019 | < 0.12 |
| c1 | Exposure preART | 0.10 | 0.04 | 2.36 | 0.019 | < 0.12 |

| Clinical variable | Biochemicals | estimate | std. error | t value | p-value | p-value adj |
| --- | --- | --- | --- | --- | --- | --- |
| ART_PI | DAG(14:0/18:3) | 0.25 | 0.03 | 8.02 | 9.51e-14 | < 1 e-6 |
| VAT | **TAG(52:2)-FA(16:0)** | **22.86** | **4.72** | **4.85** | **3e-06** | **< 0.01** |
| VAT | **TAG(52:2)-FA(18:1)** | **22.43** | **4.75** | **4.72** | **4e-06** | **< 0.01** |
| VAT | **glutamate** | **20.04** | **4.44** | **4.51** | **1.1e-5** | **< 0.01** |
| VAT | TAG(50:3)-FA(16:0) | 21.39 | 4.75 | 4.5 | 1.2e-5 | < 0.01 |
| VAT | TAG(54:4)-FA(20:4) | 21.04 | 4.63 | 4.54 | 1e-05 | < 0.01 |
| VAT | DAG(18:0/18:1) | 20.45 | 4.58 | 4.47 | 1.3e-5 | < 0.01 |
| VAT | TAG(56:4)-FA(22:4) | 20.59 | 4.62 | 4.45 | 1.4e-5 | < 0.01 |
| VAT | TAG(56:5)-FA(22:4) | 20.6 | 4.62 | 4.46 | 1.4e-5 | < 0.01 |
| VAT | TAG(54:5)-FA(22:5) | 20.03 | 4.62 | 4.33 | 2.3e-5 | < 0.01 |
| VAT | **DAG(16:0/18:1)** | **20.44** | **4.75** | **4.31** | **2.6e-5** | **< 0.01** |
| VAT | TAG(50:4)-FA(16:0) | 20.05 | 4.72 | 4.25 | 3.3e-5 | < 0.01 |
| VAT | TAG(50:2)-FA(16:0) | 20.09 | 4.78 | 4.2 | 4e-05 | < 0.01 |
| VAT | DAG(16:0/20:4) | 19.64 | 4.7 | 4.18 | 4.4e-5 | < 0.01 |
| VAT | TAG(51:3)-FA(16:0) | 19.45 | 4.72 | 4.12 | 5.6e-5 | < 0.01 |
| VAT | TAG(52:1)-FA(18:1) | 19.04 | 4.62 | 4.12 | 5.6e-5 | < 0.01 |
| VAT | TAG(50:2)-FA(18:2) | 19.11 | 4.68 | 4.08 | 6.6e-5 | < 0.01 |
| VAT | TAG(50:3)-FA(18:3) | 19.05 | 4.67 | 4.08 | 6.4e-5 | < 0.01 |
| VAT | TAG(51:4)-FA(16:0) | 19.34 | 4.73 | 4.08 | 6.4e-5 | < 0.01 |
| VAT | **TAG(54:3)-FA(20:3)** | **18.91** | **4.71** | **4.01** | **8.6e-5** | **< 0.01** |
| VAT | TAG(50:1)-FA(16:0) | 18.68 | 4.72 | 3.96 | 1.06e-4 | < 0.07 |
| VAT | TAG(52:1)-FA(16:0) | 18.23 | 4.6 | 3.96 | 1.06e-4 | < 0.07 |
| VAT | TAG(50:3)-FA(18:2) | 18.86 | 4.79 | 3.94 | 1.13e-4 | < 0.07 |
| VAT | TAG(54:4)-FA(22:4) | 18.09 | 4.63 | 3.9 | 1.3e-4 | < 0.07 |
| VAT | TAG(50:1)-FA(18:1) | 17.92 | 4.72 | 3.79 | 1.98e-4 | < 0.07 |
| VAT | TAG(52:2)-FA(18:0) | 17.43 | 4.59 | 3.8 | 1.95e-4 | < 0.07 |
| ART_PI | PI(16:0/20:3) | 0.13 | 0.03 | 3.74 | 2.43e-4 | < 0.07 |
|  | Continued on next page | | |  |  |  |

**Supplementary Table S5:** Association table predicting clinical variables based on each lipid and metabolite concentration (FDR < 0.07) within community c1. The model is adjusted for MetS, sex and age. Ranked according to FDR (p-value adj). Five key lipids and one key metabolite were associated with VAT and marked in bold in the table.

| Clinical variable | Biochemicals | estimate | std. error | t value | p-value | p-value adj |
| --- | --- | --- | --- | --- | --- | --- |
| VAT | TAG(53:4)-FA(16:0) | 17.31 | 4.68 | 3.7 | 2.83e-4 | < 0.07 |
| VAT | **TAG(54:3)-FA(20:2)** | **17.4** | **4.74** | **3.67** | **3.13e-4** | **< 0.07** |
| VAT | TAG(52:3)-FA(20:3) | 17.13 | 4.72 | 3.63 | 3.64e-4 | < 0.07 |
| VAT | TAG(52:4)-FA(20:4) | 16.51 | 4.6 | 3.59 | 4.21e-4 | < 0.07 |
| VAT | TAG(51:3)-FA(18:3) | 16.34 | 4.61 | 3.54 | 4.95e-4 | < 0.07 |
| VAT | TAG(50:2)-FA(16:1) | 16.84 | 4.76 | 3.54 | 5e-04 | < 0.07 |
| VAT | DAG(16:1/20:4) | 16.53 | 4.67 | 3.54 | 5.09e-4 | < 0.07 |
| VAT | TAG(52:1)-FA(18:0) | 15.98 | 4.55 | 3.52 | 5.46e-4 | < 0.07 |
| VAT | DAG(16:1/18:0) | 16.17 | 4.63 | 3.49 | 5.91e-4 | < 0.07 |
| VAT | TAG(51:2)-FA(16:0) | 16.44 | 4.73 | 3.47 | 6.32e-4 | < 0.07 |
| VAT | TAG(50:2)-FA(18:1) | 16.55 | 4.79 | 3.45 | 6.79e-4 | < 0.07 |
| VAT | TAG(53:3)-FA(16:0) | 15.82 | 4.63 | 3.42 | 7.75e-4 | < 0.07 |
| VAT | DAG(16:0/16:1) | 15.97 | 4.73 | 3.37 | 8.98e-4 | < 0.07 |
| VAT | TAG(51:3)-FA(17:0) | 15.88 | 4.71 | 3.37 | 8.98e-4 | < 0.07 |
| VAT | TAG(53:2)-FA(17:0) | 15.54 | 4.6 | 3.37 | 8.91e-4 | < 0.07 |
| VAT | TAG(54:5)-FA(22:4) | 15.92 | 4.72 | 3.37 | 8.99e-4 | < 0.07 |
| VAT | TAG(52:5)-FA(20:4) | 15.7 | 4.67 | 3.36 | 9.46e-4 | < 0.07 |
| VAT | TAG(50:1)-FA(16:1) | 15.49 | 4.65 | 3.33 | 0.001044 | < 0.07 |
| SAT | TAG(48:4)-FA(18:1) | -16.1 | 4.9 | -3.29 | 0.001196 | < 0.07 |
| VAT | TAG(54:2)-FA(20:2) | 15.33 | 4.66 | 3.29 | 0.0012 | < 0.07 |
| ART_NNRTI | TAG(48:2)-FA(12:0) | 0.12 | 0.04 | 3.28 | 0.00123 | < 0.07 |
| VAT | TAG(53:4)-FA(20:4) | 15.17 | 4.63 | 3.28 | 0.001238 | < 0.07 |
| VAT | DAG(16:0/18:0) | 14.99 | 4.58 | 3.27 | 0.001258 | < 0.07 |
| VAT | TAG(52:4)-FA(22:4) | 15.11 | 4.62 | 3.27 | 0.00126 | < 0.07 |
| VAT | TAG(51:2)-FA(18:2) | 15.04 | 4.61 | 3.26 | 0.001304 | < 0.07 |
| VAT | TAG(53:5)-FA(20:4) | 15.18 | 4.68 | 3.24 | 0.001385 | < 0.07 |
| ART_NNRTI | TAG(44:2)-FA(18:1) | 0.12 | 0.04 | 3.23 | 0.001444 | < 0.07 |
| VAT | DAG(16:1/18:1) | 15.36 | 4.75 | 3.23 | 0.001448 | < 0.07 |
| VAT | DAG(16:0/20:3) | 15.36 | 4.76 | 3.23 | 0.001456 | < 0.07 |
| VAT | TAG(52:6)-FA(20:4) | 15.19 | 4.71 | 3.23 | 0.001475 | < 0.07 |
| VAT | DAG(14:0/18:1) | 14.97 | 4.66 | 3.21 | 0.001551 | < 0.07 |
| VAT | TAG(48:3)-FA(16:0) | 14.89 | 4.64 | 3.21 | 0.001563 | < 0.07 |
| VAT | TAG(50:3)-FA(16:1) | 15.1 | 4.75 | 3.18 | 0.00171 | < 0.07 |
| VAT | TAG(50:2)-FA(14:0) | 14.77 | 4.66 | 3.17 | 0.001786 | < 0.07 |
| ART_NNRTI | CER(14:0) | 0.11 | 0.04 | 3.16 | 0.001843 | < 0.07 |
| ART_NNRTI | TAG(42:1)-FA(18:1) | 0.11 | 0.03 | 3.15 | 0.001879 | < 0.07 |
| VAT | TAG(48:2)-FA(18:2) | 14.38 | 4.57 | 3.15 | 0.001896 | < 0.07 |
| VAT | TAG(51:1)-FA(17:0) | 14.49 | 4.6 | 3.15 | 0.001913 | < 0.07 |
| VAT | TAG(48:3)-FA(14:0) | 14.73 | 4.69 | 3.14 | 0.001945 | < 0.07 |
|  | Continued on next page | | |  |  |  |

| Clinical variable | Biochemicals | estimate | std. error | t value | p-value | p-value adj |
| --- | --- | --- | --- | --- | --- | --- |
| VAT | TAG(50:0)-FA(16:0) | 14.3 | 4.55 | 3.14 | 0.001937 | < 0.07 |
| VAT | TAG(52:2)-FA(16:1) | 14.41 | 4.6 | 3.13 | 0.002008 | < 0.07 |
| VAT | TAG(51:2)-FA(15:0) | 14.51 | 4.64 | 3.13 | 0.00203 | < 0.07 |
| ART_NNRTI | TAG(44:1)-FA(18:1) | 0.11 | 0.04 | 3.12 | 0.002072 | < 0.07 |

| Rank | Biochemicals | Degree |
| --- | --- | --- |
| 1 | TAG(50:3)-FA(18:2) | 639 |
| 2 | TAG(51:3)-FA(17:0) | 637 |
| 3 | DAG(18:0/18:1) | 636 |
| 4 | TAG(50:3)-FA(16:0) | 636 |
| 5 | TAG(50:4)-FA(16:0) | 636 |
| 6 | TAG(53:2)-FA(17:0) | 635 |
| 7 | TAG(51:3)-FA(16:0) | 634 |
| 8 | TAG(53:4)-FA(16:0) | 633 |
| 9 | TAG(52:2)-FA(20:0) | 632 |
| 10 | TAG(51:3)-FA(16:1) | 631 |
| 11 | TAG(48:4)-FA(16:1) | 630 |
| 12 | TAG(50:2)-FA(14:0) | 630 |
| 13 | **TAG(52:2)-FA(16:0)** | **630** |
| 14 | TAG(52:2)-FA(16:1) | 630 |
| 15 | **TAG(52:2)-FA(18:1)** | **630** |
| 16 | TAG(50:3)-FA(18:1) | 629 |
| 17 | TAG(51:4)-FA(16:0) | 629 |
| 18 | TAG(52:4)-FA(14:0) | 629 |
| 19 | TAG(53:2)-FA(18:1) | 629 |
| 20 | TAG(54:5)-FA(22:5) | 629 |
| 21 | DAG(14:0/18:2) | 628 |
| 22 | TAG(51:2)-FA(18:1) | 628 |
| 23 | TAG(52:5)-FA(14:0) | 628 |
| 24 | TAG(54:4)-FA(20:4) | 628 |
| 25 | TAG(52:3)-FA(20:2) | 627 |
| 26 | TAG(50:2)-FA(18:1) | 627 |
| 27 | TAG(52:4)-FA(20:2) | 627 |
| 28 | TAG(52:5)-FA(20:3) | 627 |
| 29 | TAG(50:3)-FA(14:1) | 626 |
| 30 | TAG(53:5)-FA(20:4) | 626 |
| 31 | TAG(52:6)-FA(20:4) | 625 |
| 32 | **TAG(54:3)-FA(20:3)** | **625** |
| 33 | DAG(16:0/20:4) | 624 |
| 34 | TAG(52:7)-FA(18:1) | 624 |

**Supplementary Table S6:** Top 10% nodes in community c1 based on the degree. Thus, the most interconnected nodes in the community. The lipids TAG(52:2)-FA(16:0), TAG(52:2)-FA(18:1) and TAG(54:3)-FA(20:3) are marked in bold, as they were among the key lipids.

Supplementary Figures

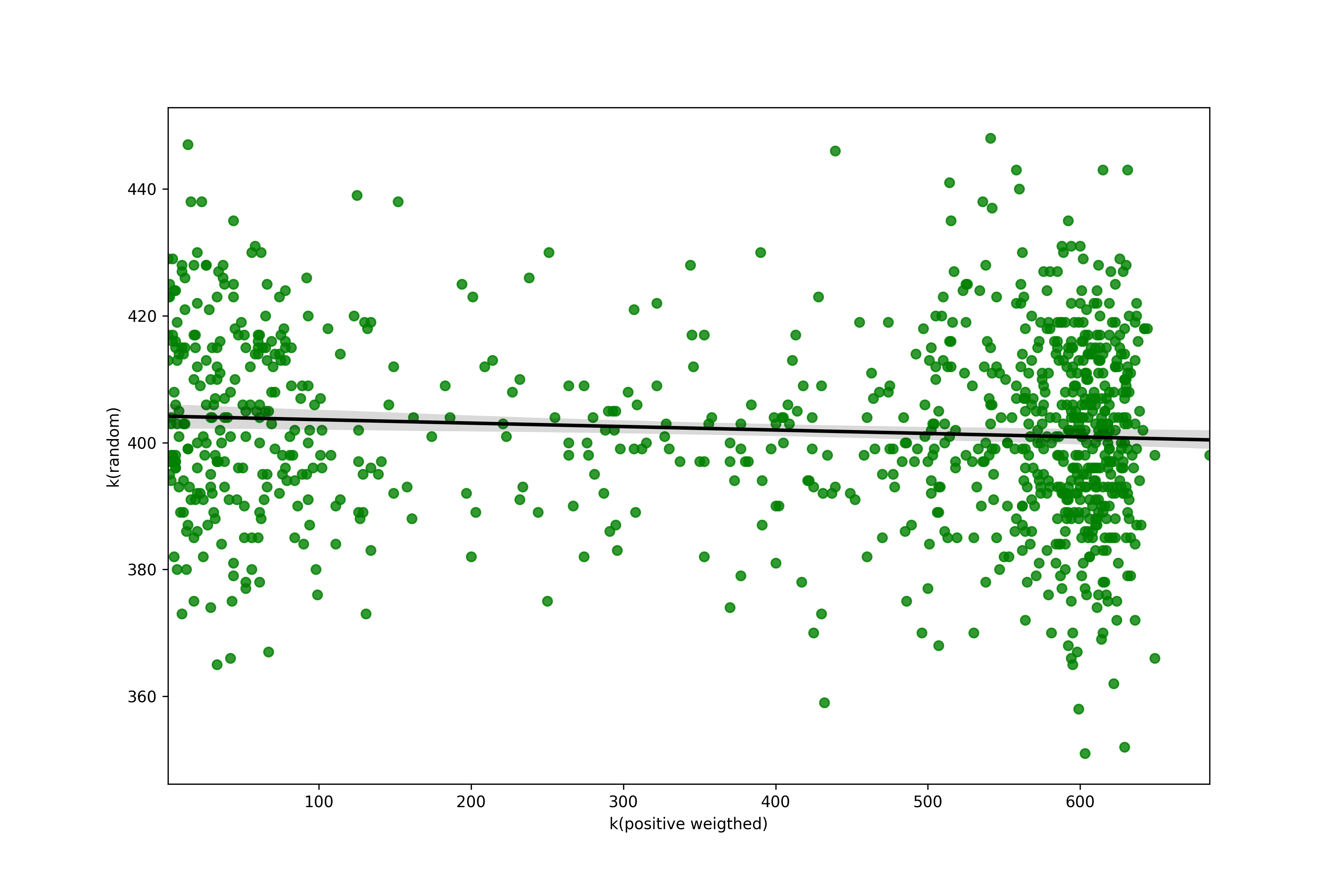

**Supplementary Figure S1:** Degree distribution for the positive weighted against the random network.

**a**.
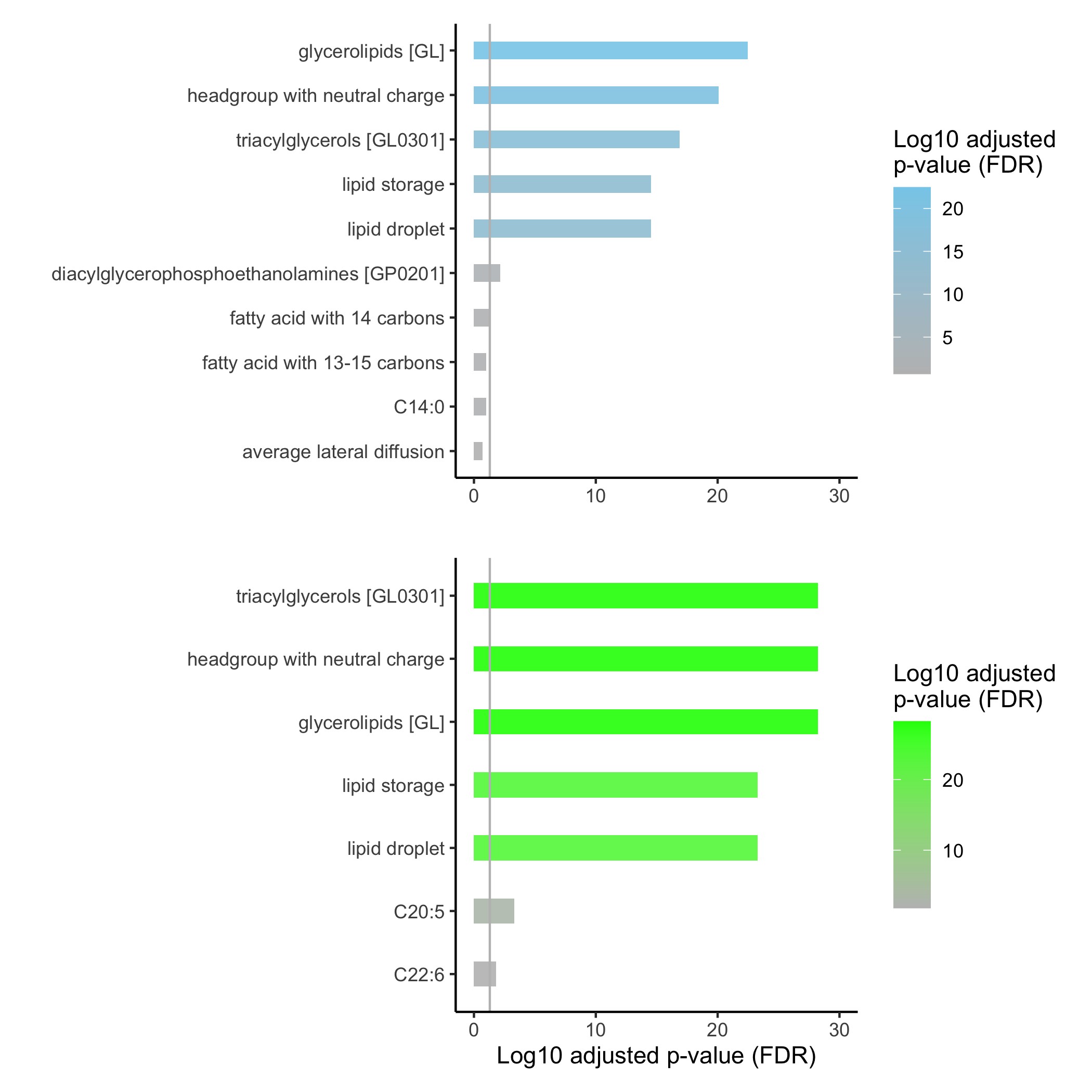

**b.**
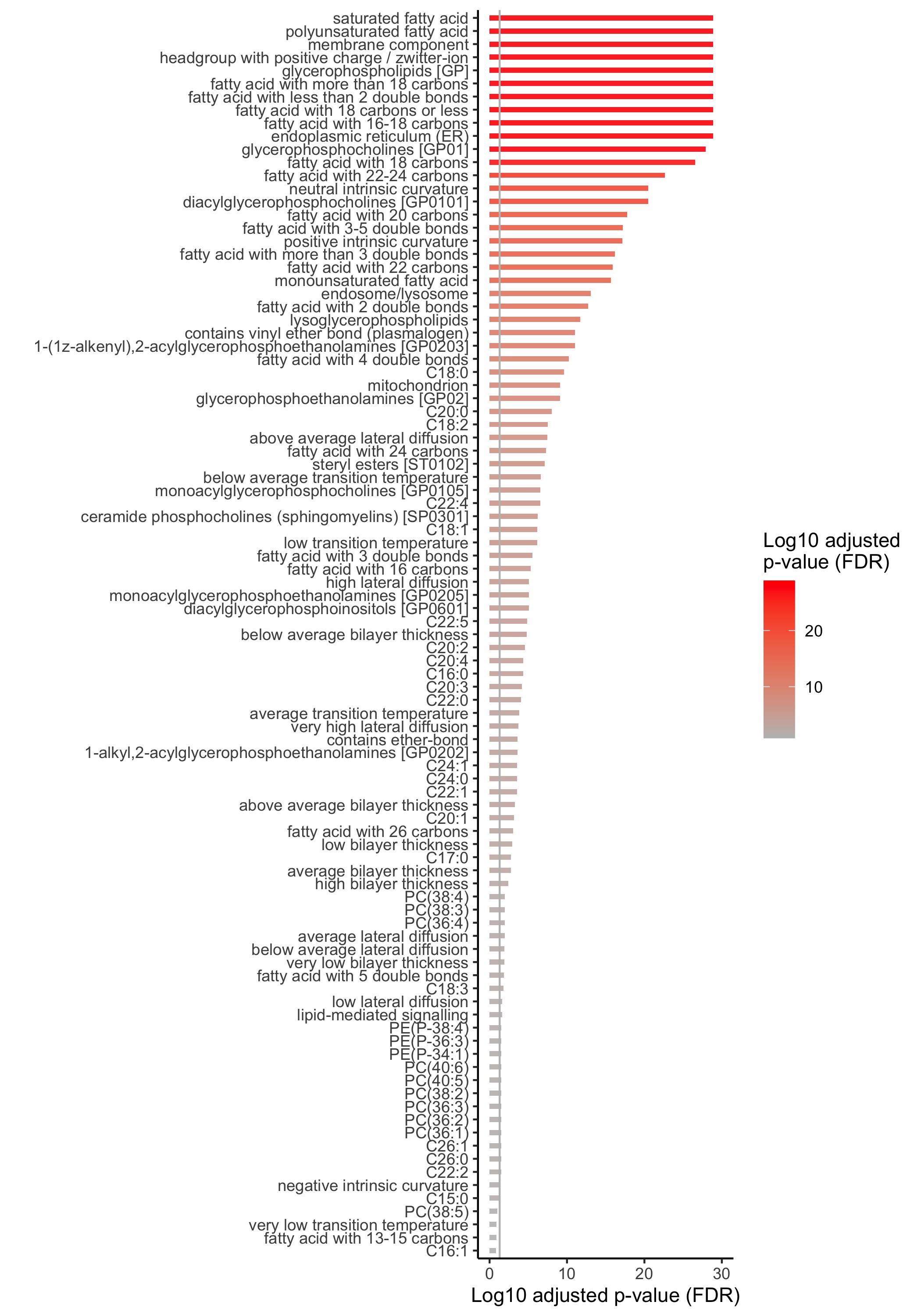

**Supplementary Figure S2:** Ontology enrichment plots for the three communities c1, c2 and c3. (a) Ontology enrichment plots for community c1 (blue) and c2 (green). (b) Ontology enrichment plot for community c3.
